# Impact of a Multidisciplinary Nutritional Support Team on Quality Improvement for Patients Receiving Home Parenteral Nutrition

**DOI:** 10.1101/2022.04.06.22273454

**Authors:** Michael M. Rothkopf, Mohan D. Pant, Rebecca Brown, Jamie Haselhorst, Francine Gagliardotto, Allison Tallman, Debbie Stevenson, Andrew DePalma, Michael Saracco, Dan Rosenberg, Vladimir Proudan, Kishwar Shareef, Nudrat Ayub

## Abstract

**Introduction:** Home parenteral nutrition (HPN) is an essential therapy for patients requiring long term nutritional support. The Amerita Quality Improvement Project for HPN Patients (QIP-PN) explored opportunities for QI for patients under its service. As a component of QIP-PN,we studied the effect of a Physician Nutrition Expert (PNE)-led multidisciplinary nutritional support team (MNST) on HPN care.

**Objective:** To test the effect of an MNST on adherence to protocols, outcomes and QOL in HPN.

**Methods:** The study was divided into 3 phases: data review (phases 1a and 1b), observation (phase 2) and intervention (phase 3). 7 Amerita branch locations were selected as “study branches” based upon their volume of long-term HPN cases. All patients in the study were drawn from this population. Since the study was part of a QI project rather than a randomized controlled study, we employed a quasi-experimental design with a case-matched control group (control). Data were collected on demographics, treating physicians PNE status, HPN care variables, recommended interventions, quality-of-life assessment, adverse outcomes and hospitalizations. Paired t-test was used to compare continuous data between phases 2 and 3. Comparison between the study and control groups utilized a negative binomial regression model. Statistical analysis utilized R (https://www.r-project.org/).

**Results:** 34 patients were reviewed in phase 1a and 197 in phase 1b . 40 study patients completed phase 2 and progressed into phase 3, of whom 30 completed ≥60 therapy days. Improvements in weight, BMI and QOL were seen in the study patients during intervention. Recommendations made and accepted by treating physicians differed based on PNE status. Study patients had fewer adverse outcomes and related hospitalizations than controls.

**Conclusion:** MNST recommendations improved clinical, biochemical parameters and patients’ self-reported overall health. MNST input reduced adverse outcomes, hospitalization and hospital length of stay. This study highlights the potential for MNST to have a significant impact on the quality and overall cost of HPN management.

## Introduction

Home parenteral nutrition (HPN) is a life-sustaining therapy for more than 25,000 patients in the US alone (79 patients/million inhabitants) (1-3). HPN permits these individuals to continue functional lifestyles without hospital confinement, performing self-care or receiving assistance from family members (4-5). Home infusion reduces the cost of caring for PN-dependent patients by as much as 65% (6).

The known risks of HPN (catheter-related bloodstream infections; CRBSIs, venous thrombosis, metabolic imbalances, bone disease, kidney stones, and liver disease) often lead to emergency department visits and hospital re-admission (7-11) and the burden of performing home infusion therapy can impact patients’ quality of life (QOL) (12-15). Therefore, most US patients are managed by a home infusion provider and closely monitored by a homecare team. The infusion provider supplies the PN formulation, infusion pump, tubing, dressings, etc. to facilitate the homecare infusion process. A homecare nurse visits regularly to examine the patient, record vital signs, check the catheter site and obtain specimens for monitoring.

The HPN nutrition support team (NST) is typically comprised of homecare specialists in pharmacology, nutrition, and nursing. At centers like the Mayo Clinic, NSTs may include physician nutrition experts (PNE) (16). In most instances, home infusion providers coordinate HPN with the patient’s physician. The input of a PNE is not a requirement for home infusion companies to provide HPN (17).

Utilization of HPN standards often depends upon the expertise of the treating physicians (18-20). This is concerning because our internal data shows that the majority of HPN patients are managed by physicians without nutrition certification (21).

Despite widespread HPN use, data is lacking on both the adherence to established protocols and the QOL of HPN consumers (22-25). This investigator-initiated study explored opportunities for quality improvement (QI) in HPN. Our objective was to test the effect of a multidisciplinary NST (MNST), which included PNE input, on adherence to protocols, outcomes of HPN care variables and QOL in HPN. The primary hypothesis was that MNST intervention would improve HPN care variables. Another hypothesis was that the MNST intervention would decrease adverse events such as unplanned hospitalizations. In this report, we detail our findings on a QI project for HPN management guided by a MNST.

## Methods

A quality improvement project for HPN patients (QIP-PN) was established by Amerita, Inc., a national home infusion organization (Amerita). A MNST was created, consisting of a PNE, certified nutrition support clinicians (CNSC; RD, RN, RPh) and administrators. A QIP-PN study protocol was developed to examine multiple aspects of care for all HPN patients serviced by the organization. The protocol was granted Institutional Review Board (IRB) exemption under NIH guidelines (45CFR 46.104(d)(2)) by the Western IRB on September 4, 2019. A study Oversight and Safety Committee was established consisting of three non-affiliated PNEs.

### Study Design

The study was divided into 3 phases: data review (phases 1a and 1b), observation (phase 2) and intervention (phase 3) (Figure 1). Seven Amerita branch locations were selected as “study branches” based upon their volume of long-term HPN (>90 days of therapy) cases and all patients in the study were drawn from this population. Since the study was part of a QI project rather than a randomized controlled study, we employed a quasi-experimental design with a case-matched control group.

**Figure 1.**
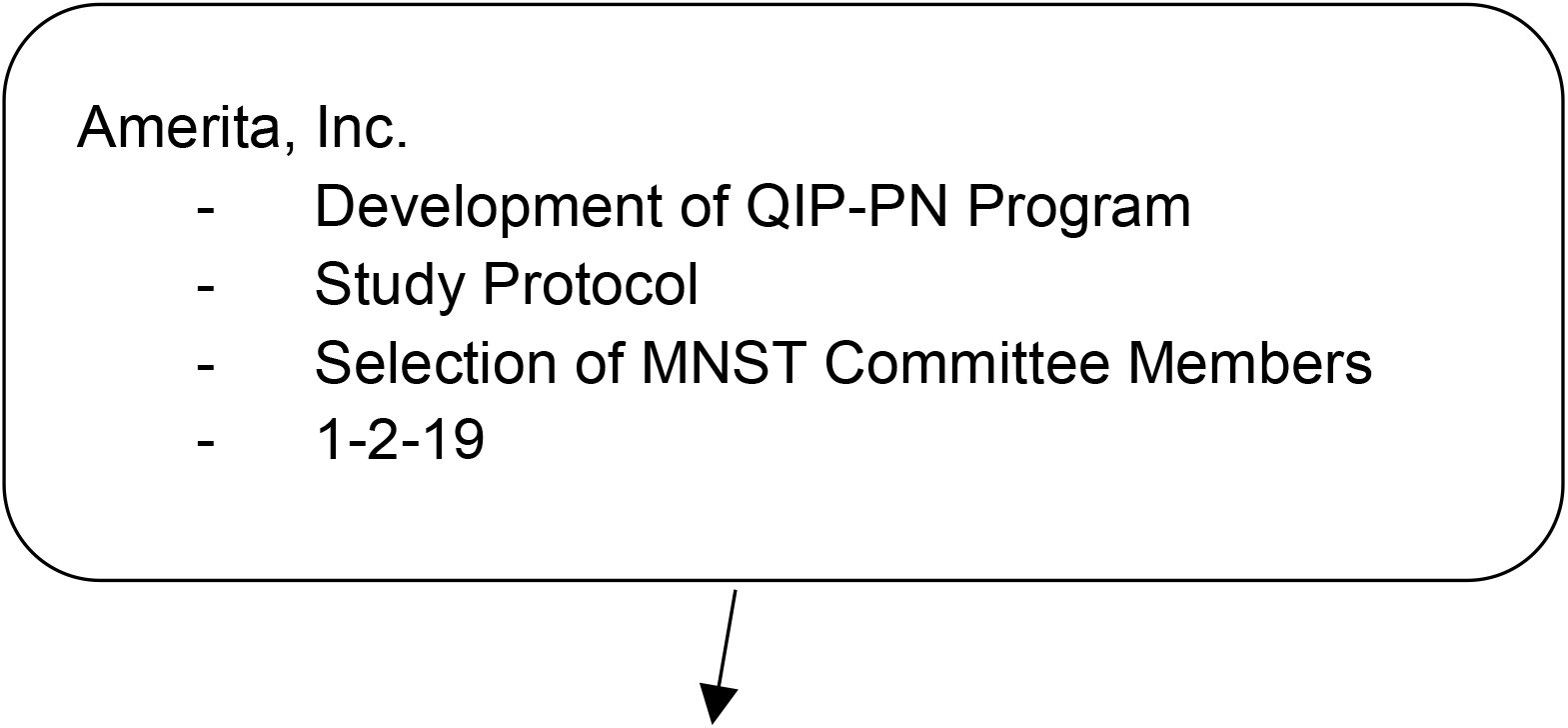
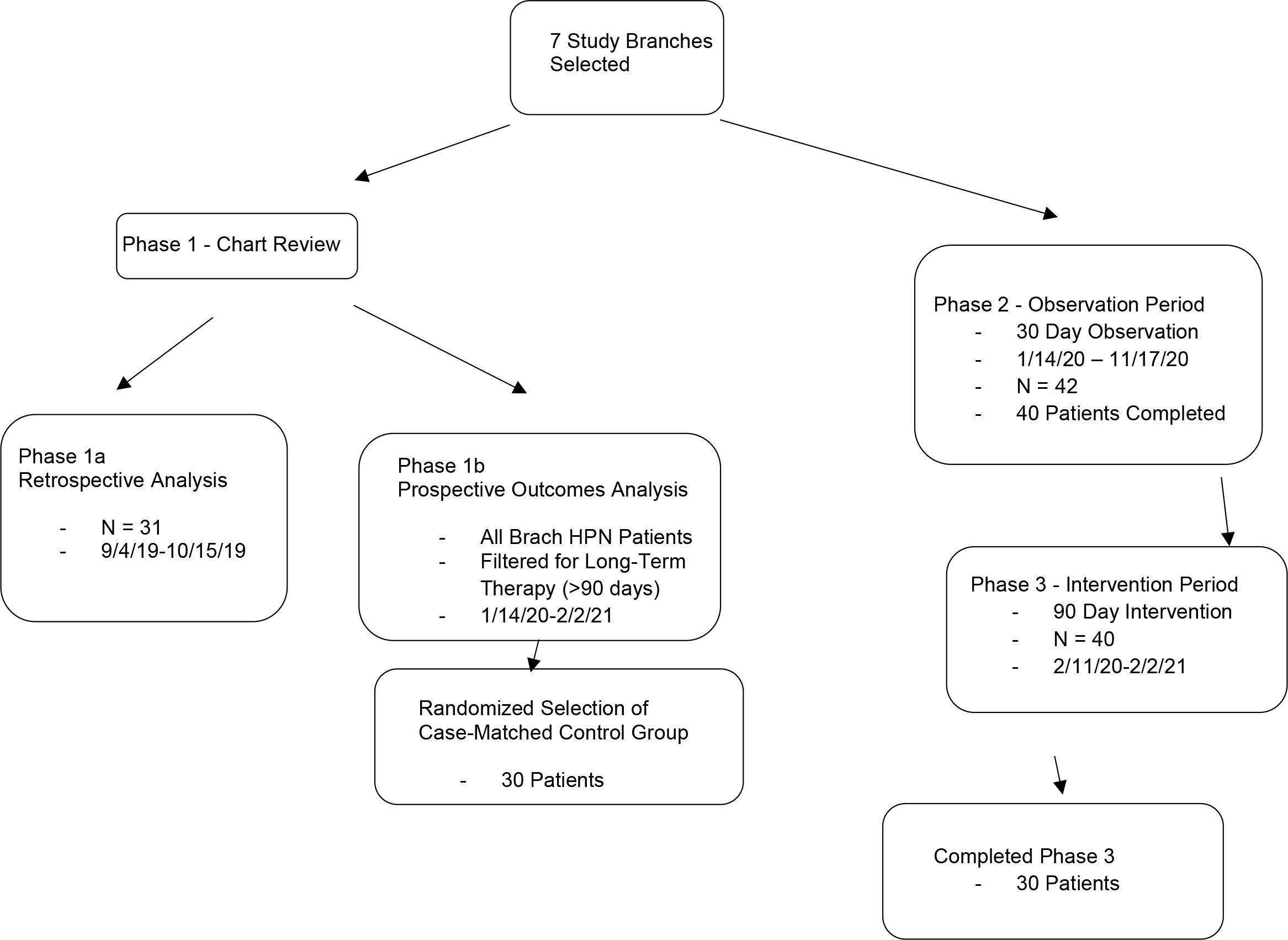
Flowsheet of patient selection and participation in the Amerita QIP-PN Study

### Study groups

Please note that the actual number of patients in any group varied because patients who were selected may have come off service before data collection was complete. Thirty (30) or more long-term HPN patients were randomly selected from the study branches for Phase 1a. Phase 1a was solely intended to confirm that the study parameters (Table 1) could be retrieved from Amerita patient records. All long-term HPN patients treated at study branches were included in Phase 1b. Patients who went on to participate in Phases 2 and 3 were removed from the Phase 1b database. All study branch long term PN patients were offered participation in phases 2 and 3. After a patient gave informed consent, their PN prescribing physician was asked to sign physician study participation agreements. Thirty (30) or more patients in whom both a signed informed consent and signed treating physician study participation agreement was obtained comprised the study group (Figure 1). The case-matched control group patients (controls) were randomly selected from long-term HPN patients from the 7 study branches. The control patients had similar referral source hospitals, percentage of PNE treating physicians, insurance coverage and demographics to the study group.

**Table 1.**
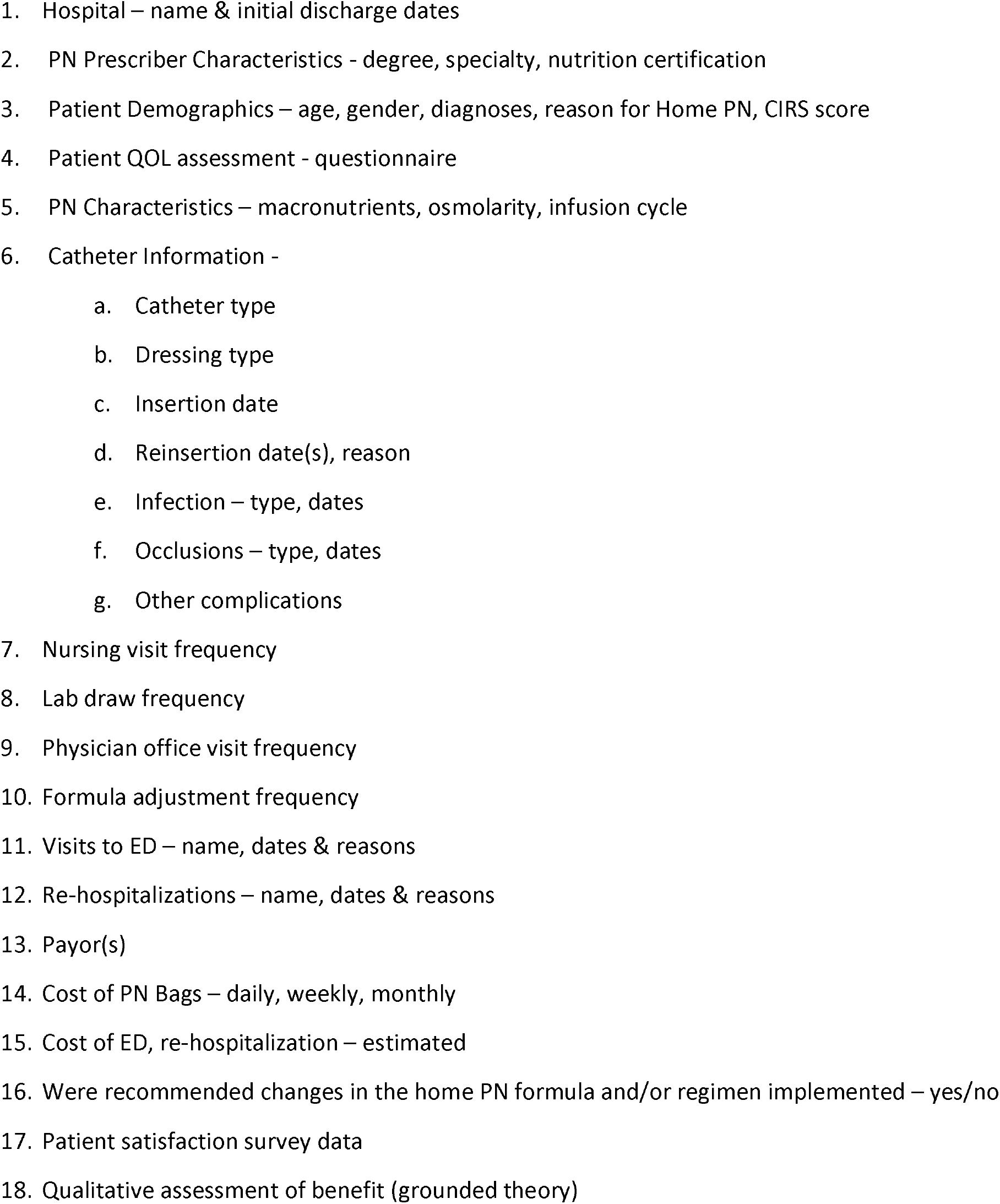
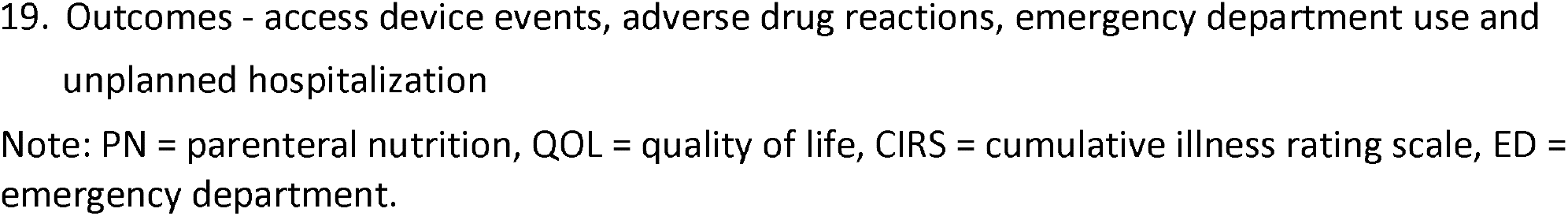
Study parameters examined in patients during phases 1a, 2 and 3.

### Study Components and Timing

Phase 1a was a 90-day retrospective review of patient data, conducted from September 4, 2019 through October 15, 2019. Phase 1b was a prospective analysis of 4 outcome measures (access device events, adverse drug reactions, emergency department use and unplanned hospitalization) of all study branch patients, conducted from January 14, 2020 through February 2, 2021. Phase 2 was a 30-day observation period of the study group patients performed between January 14, 2020 and November 17, 2020. Phase 3 was a 60-90 day review and recommendation period of each patient in the study group, conducted from February 11, 2020 through February 2, 2021. During Phase 3, the MNST made management recommendations for adjustments in HPN care in accordance with established practice guidelines. These recommendations for HPN care modification were provided to the patient’s treating physician by an Amerita branch clinician. The acceptance rate of MNST recommendations was recorded.

### Data Collection

In phase 1b, outcomes on access device events, adverse drug reactions, emergency department visits and unplanned hospitalization were monitored prospectively in study branch patients as part of the National Home Infusion Foundation (NHIF) benchmark reporting process. In phases 2 and 3, data on the study parameters (Table 1) were prospectively collected on enrolled study patients during weekly virtual meetings of the MNST.

### Statistical analysis

For the comparison between the phase 3 study group and controls, an independent samples t-test was used only when the outcome variable is assumed to have normal distribution in the population. Comparisons between phase 2 and phase 3 patient data were performed by paired t-test. This was possible because the same group of patients were used in both phases. Therefore, the study patients in the observation period (phase 2) served as their own controls for the intervention study (phase 3). Comparison between the study and control groups utilized a negative binomial regression model for modeling outcome variable, rate of adverse events per 90-day period. A negative binomial regression model was needed for modeling rate of adverse events per 90-day period as it is a count variable. Statistical analysis was conducted using R (https://www.r-project.org/).

### PNE Status of treating physicians

HPN treating physicians were classified as being either PNE or non-PNE. PNE was defined as those physicians who were either board certified by the National Board of Physician Nutrition Specialists (NBPNS) or had CNSC designation.

### Study instruments

#### Patient QOL assessment

The Euroquol 5 dimension, 3 level (EQ-5D-3L) quality-of-life instrument was chosen for its broad acceptance in the literature and prior application in HPN (31-33). The EQ-5D-3L system is comprised of five dimensions: mobility, self-care, usual activities, pain/discomfort and anxiety/depression. Each dimension has 3 levels: no problems, some problems, and extreme problems. The patient is asked to indicate their health state by selecting the choice corresponding to the most appropriate statement in each of the five dimensions. This decision results in a 1-digit number that expresses the level selected for that dimension. A sixth dimension records the patient’s overall self-rated health on a 0-100 scale where the endpoints are labelled ‘Best imaginable health state’ and ‘Worst imaginable health state’. The overall health state is a quantitative measure that reflects the patient’s own perception of health. This component is scored as a visual analog scale (VAS).

The EQ-5D-3L was administered at the start of phase 2, start and end of phase 3. Due to COVID-19 restrictions on in-person contact, the EQ-5D-3L was administered via a telephone interview by an independent patient care coordinator who was not a member of the MNST. Such telephonic methodology is in accordance with the EQ-5D-3L guidelines (34). For this study we used VAS score as a proxy for QOL.

#### Measure of multi-morbidity and disease burden

We explored the use of multimorbidity scales for their application to HPN patients (35) and elected to measure disease burden on study and control patients by means of the Cumulative Illness Rating Scale (CIRS) (36). This method analyzes the multimorbidity of an individual patient by reviewing 14 body system categories which are graded from zero to four (37-39). The CIRS approach has been validated by numerous studies in the literature. This index can measure a chronic medical illness burden in individual patients and has been validated as a predictor of hospitalization and readmission for older adults and as a predictor of long-term mortality. The CIRS score can range from 0 to 56. Scores higher than 29 are associated with mortality during hospitalization.

Members of the MNST calculated co-morbidity and illness severity, using the CIRS, based on information available in each patient’s Electronic Medical Records (EMR) chart. This included admission notes, medications, nursing notes, emergency department notes, imaging studies, physical and occupational therapy assessments, treatment/care plans, and discharge summaries.

## Results

### Phase 1a/1b results

There were 34 patients in phase 1a. The average age was 54.7 years. Twenty-one (21) of the patients were female, 13 were male. There were 197 patients in phase 1b. These patients had 203 outcomes reported during the study period. Results for the comparison group in each outcome category were expressed in events per 1,000 days of homecare service (the NHIF standard) and events per patient (table 2).

**Table 2.** Phase 1b results.

| Adverse Outcome Event | Events/1000 HPN Days of Service | Events/Patient |
| --- | --- | --- |
| Access Device Events | 0.384 | 0.09 |
| Adverse Drug Reactions | 0.021 | 0.07 |
| Emergency Department Visits | 0.277 | 0.01 |
| Unplanned Hospitalizations | 3.647 | 0.87 |
| Therapy Related Unplanned Hospitalizations | 0.789 | 0.19 |
| Therapy Unrelated Unplanned Hospitalizations | 2.858 | 0.68 |
| Overall Adverse Outcomes | 4.33 | 1.03 |

Note. Adverse outcomes in 197 long-term HPN patients at 7 study branches.

### Phase 2/3 results

There were 42 patients enrolled in the study. Forty (40) completed 30 days or more of observation in phase 2. Thirty (30) of the 40 phase 2 patients completed 60 days or more in phase 3.

#### A. Results among participants between phases 2 and 3 for monitored parameters (Tables 3-6)

1. Patient weight, BMI and percentage of ideal body weight (%IBW) (Table 3). Average weight increased slightly between phases 2 and 3. However, 3 of the phase 3 study patients were outliers which altered these results. One was non-compliant to therapy. The other two were obese and sought to lose weight while on parenteral nutrition (PN). All other patients sought weight gain or weight stabilization. After excluding data for the 3 outliers, the study group had a statistically significant increases in weight, BMI and %IBW (calculated using the Hamwi method). Weight increased 1.78 kg; *t* (df = 26) = -3.88, *p*= .0006. BMI increased 0.7 kg/m^2^ ; *t* (df = 26) = -3.88, *p* = 0.0006). The % IBW increased by 3.66%; *t* (df = 26) = -2.26, *p* = 0.0009).
2. Basic lab results (CBC, CMP, Magnesium, Phosphate, Triglycerides). Basic lab tests were performed on a regular basis for each patient. Twenty-one (21) of the 30 patients received PN lab work on a weekly schedule. Two (2) patients received lab work every 2 weeks. Seven (7) patients received lab work on a monthly basis. The frequency of lab work orders was based on the stability of the patient, but also varied by PNE status. The majority (5 of 7) of those receiving monthly PN labs were followed by a PNE.
3. Specialized lab results. Specialized lab data on micronutrient levels was obtained in 27 of the 30 phase 3 patients (90%). Micronutrient levels were obtained prior to phase 3 monitoring in 9 of the 30 patients (30%). Seven (7) of these patients were cared for by a PNE. During phase 3 monitoring, an additional 18 patients had micronutrient levels obtained. Three (3) patients did not have micronutrient levels obtained either prior to or during phase 3 monitoring.
4. PN Component Costs and Modifications (Tables 4-5). The overall cost of PN components decreased by 1.31% during phase 3. Cost changes ranged from a decrease of 26.22% to an increase of 76.28%. Costs were reduced when excess nutrients were found to be unnecessary (i.e. thiamine, ascorbic acid and zinc included in initial discharge orders). Costs increased when additional micromineral supplementation, (i.e. selenium, zinc, and other trace elements) was required. Unprecedented, industry-wide price increases in specific components confounded MNST interventions to reduce PN costs. Product availability and shortages also impacted cost. Macronutrient orders were changed during the course of phase 3 intervention in 21 of 30 patients (70%), either because of inadequate weight gain, liver enzyme elevations or excessive weight gain. Electrolytes and micronutrient orders were changed based on lab results.
5. Patient QOL assessment (Figure 3). The average EQ-5D-3L VAS score of overall health rose from 59.41 before intervention to 71.65 by the end of the intervention period. This represents an increase of 12.24 points +/-10.0, or an improvement of 20.6% (*t*(df = 28) = -4.10, *p* = 0.0003).
6. Recommended interventions (Table 6). The MNST made a total of 157 recommendations for compliance to standards of care for each study patient. The treating physicians had final authority to either accept or reject the recommendations. Recommendations were significantly lower if the treating physician was a nutritional expert (3.09 ± 1.92 vs 5.86 ± 1.89; p=0.0001). Recommendation acceptance was high (87.2%) but was significantly lower if the treating physician was a nutritional expert (2.36 ± 2.42 vs 5.28 ± 2.22 p=0.0008). Impact on outcomes from non-accepted recommendations was not assessed.

**Table 3.**
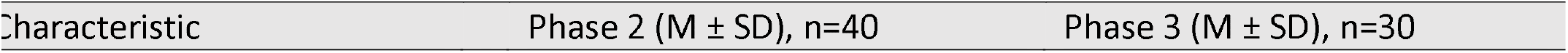
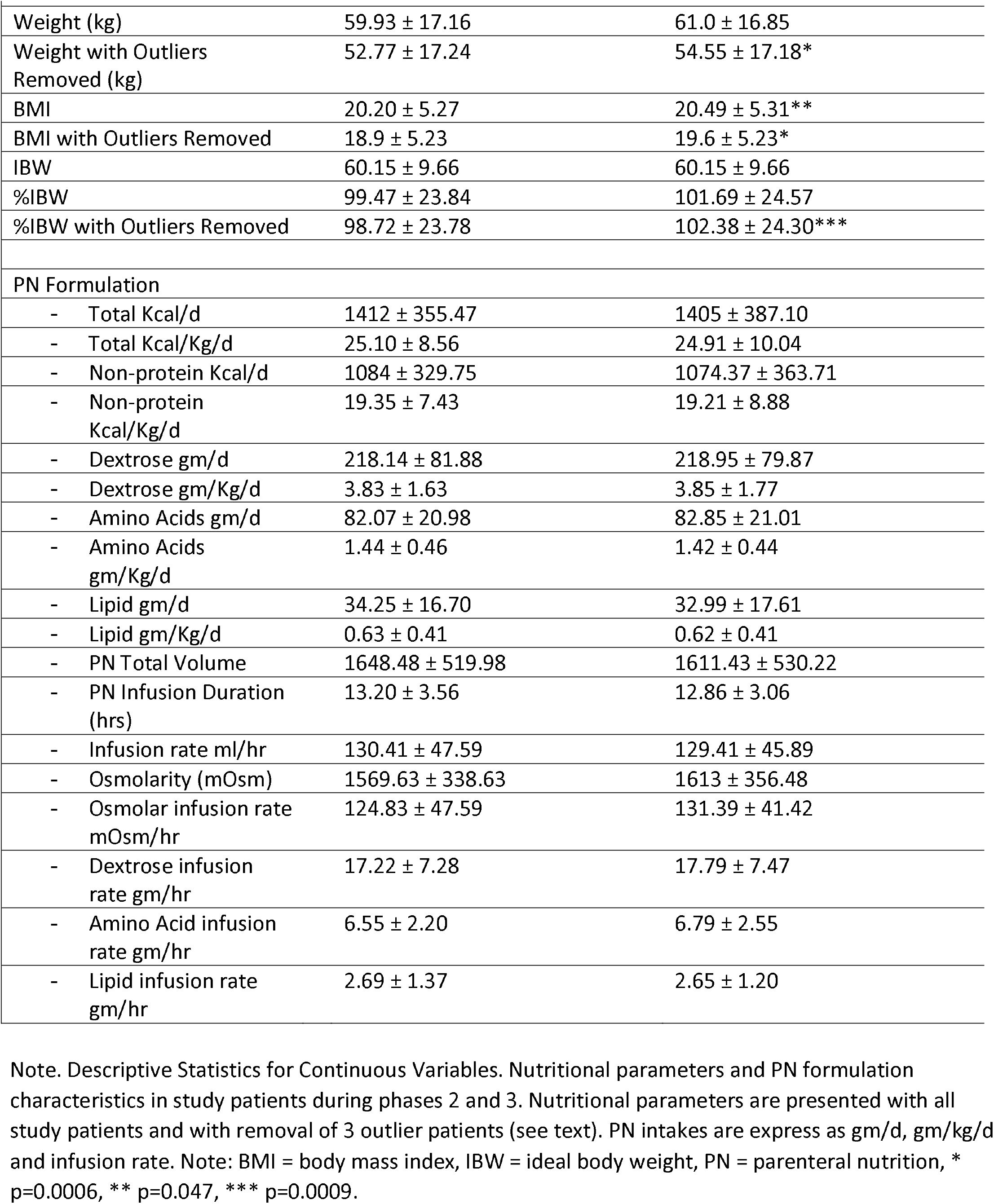
Descriptive statistics of continuous variables for patients in phases 2 and 3.

**Table 4.** Characteristic of study group patients in the context of PN cost.

| PN Formulation Cost | Number and Percentage of Study Group Patients, n= 30 |
| --- | --- |
| Decreased | 11 Patients (36.7%) |
| Unchanged | 5 Patients (16.6%) |
| Increased | 14 Patients (46.7%) |
| Range | -26.22% to +76.28% |
| Overall impact | -1.31% |

Note. Differences noted in cost of study patient PN therapy between the observation (phase 2) and intervention (phase 3). Costs were reduced when excess nutrients (i.e. thiamine, ascorbic acid and zinc) were shown to be unnecessary. Costs increased when micronutrients (i.e. selenium, zinc, and others) were deficient and required additional supplementation. Unprecedented, industry-wide price increases in PN components confounded the impact of MNST interventions intended to reduce PN costs. Note: PN = parenteral nutrition.

**Table 5.** PN order changes in study patients during phase 3.

| PN Order Changes | PN Modifications in Study Group Patients, n= 30 | Total PN Modifications |
| --- | --- | --- |
| Macronutrients | 21 (70%) | 104 |
| Electrolytes | 15 (50%) | 173 |
| Micronutrients | 25 (83.3%) | 42 |
| - Thiamine | 5 |  |
| - Ergocalciferol | 5 |  |
| - Ascorbic acid | 1 |  |
| - Cyanocobalamin | 1 |  |
| - Zinc | 10 |  |
| - Chromium | 5 |  |
| - Selenium | 5 |  |
| - Manganese | 6 |  |
| - Copper | 2 |  |
| Volume | 17 (56.6%) | 33 |
| Infusion Rate | 6 (20%) | 11 |
| Total | 27 | 373 |

Note. Macronutrient orders were changed either because of inadequate weight gain, liver enzyme elevations or excessive weight gain. Electrolytes and micronutrient orders were changed based on lab results. Note: PN = parenteral nutrition.

**Table 6.**
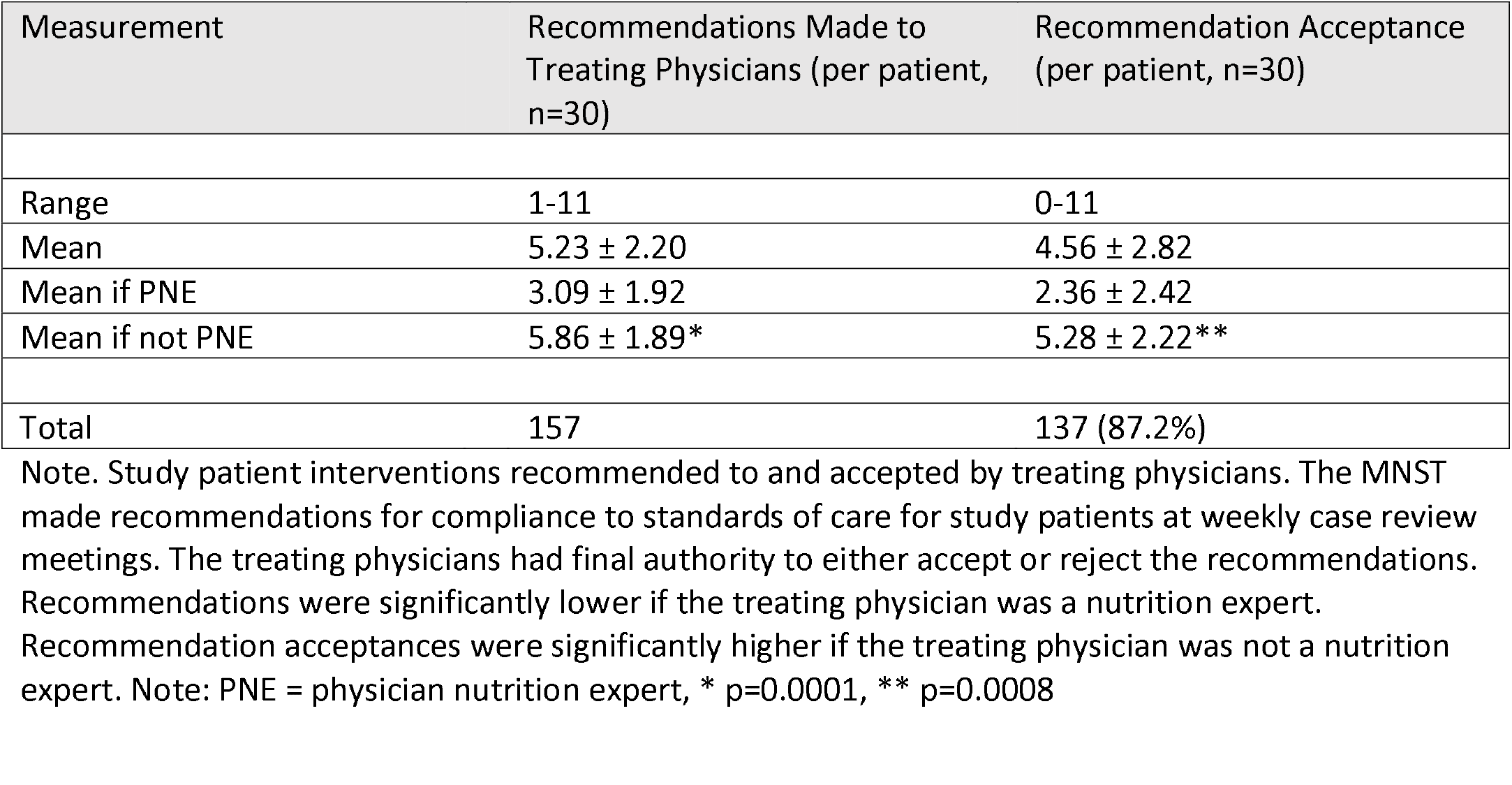
Recommendations and Acceptances by Treating Physicians.

#### B. Comparison between study patients and case-matched controls (Tables 7-11)

1. Patient characteristics (Tables 7-8). Study patients and controls had similar characteristics with regard to gender, age, length of time on HPN, primary diagnosis, comorbidity (CIRS Score) and payor mix. Hospital referral source and the minority PNE prescriber status were also similar between the groups.
2. Catheter Characteristics (Table 9). Both groups had similar numbers of infusion ports. The study group had more tunneled catheters (i.e. Hickman®, Broviac®, Groshong® catheters) whereas control patients had more peripherally inserted central catheter (PICC) lines. More study group patients were visited by Amerita staff nurses than by home health agency nurses. A similar number of patients in both groups had bio-occlusive catheter dressings.
3. PN order changes (Table 10). PN formula changes were 20% more frequent among the study patients than controls. Macronutrients were modified 142.4% more often, micronutrients 350% more often and volume 137.5% more often in study patients than controls. Conversely, electrolytes were modified 97% as often and duration 84.6% as often in study patients than controls.
4. Adverse Outcomes (Table 11). There were 10 total outcomes reported among the phase 3 patients (0.33/patient), including 7 unplanned hospitalizations. Three (3) of the hospitalizations were therapy related. There were 2 emergency department (ED) visits among the phase 3 patients during the study period. Phase 3 patients had lower total adverse outcomes compared to the data from the 197 long term HPN patients at the 7 study branches (3.64 vs 4.33), unplanned hospitalization (2.54 vs 3.65) and access device events (0.338 vs 0.432) per 1,000 therapy days. ED use was higher in the study patients (0.338 vs 0.299) per 1,000 therapy days. There were 16 total adverse outcomes among the control patients (0.53/patient), with 14 unplanned hospitalizations. Three (3) hospitalization were therapy related. There were 2 ED visits among the control patients during the study period.
5. Total Hospitalizations and Length of Stay (LOS). Study group patients were hospitalized a total of 11 times during phase 3 (hospitalization rate = 0.37 admissions/patient). The total LOS was 69 days. The average LOS (hospital days/ number of hospitalizations) for the study group was 6.27 days. Among the 7 study patients hospitalized, 3 had only one hospitalization, 4 had a second hospitalization while none had a third. The readmission rate of study group patients was 0.13 (number of readmissions/ number of patients). Control group patients were hospitalized a total of 20 times during the 90-day review (hospitalization rate of 0.67). The total LOS was 153 days. The average LOS was 7.65 days. Among the 13 control group patients who were hospitalized, 8 had only one hospitalization, 5 had a second hospitalization and 2 had a third. The readmission rate of control group patients was 0.23 (Table 11).
6. Statistical Analysis of Phase 3 Study Patients and Controls (Table 12). Comparison between the study and control group utilized a negative binomial regression model for modeling outcome variable, rate of adverse events per 90-day period, based on zero-inflated count data. We used a negative binomial regression model to explain how this outcome is related with other possible predictors and to create a prediction model. Among the available count-data based predictors, 90-day related hospitalizations, 90-day access device events, and 90-day total change were chosen as possible predictors because of their low correlation with 90-day events and with each other.

**Table 7.**
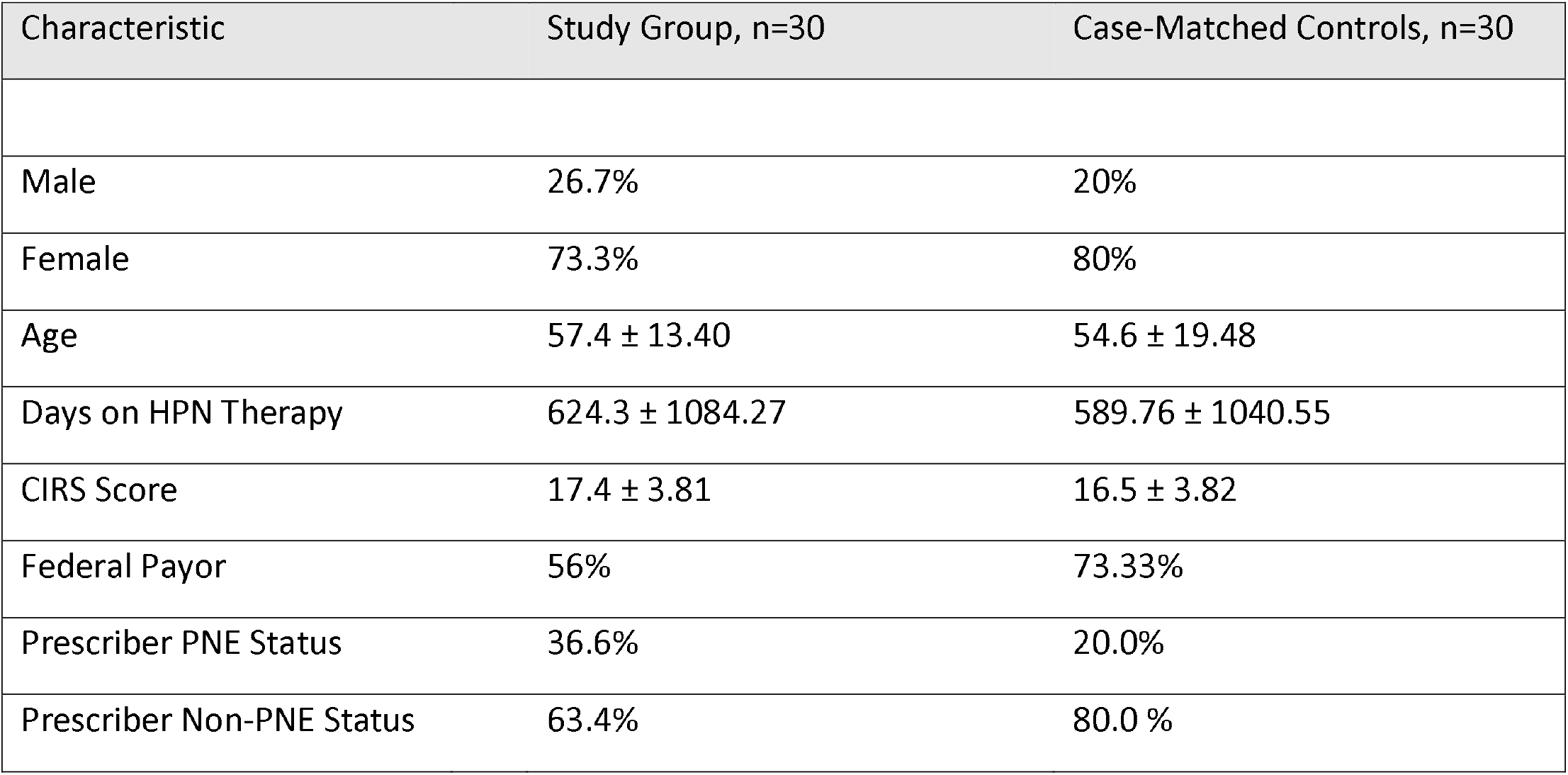
Demographic characteristics of study patients and case-matched controls. Note. No significant demographic differences were identified between the groups. Note: CIRS = cumulative illness rating scale, PNE = physician nutrition expert

**Table 8.** Diagnostic categories of study patients and controls.

| Primary Diagnosis | Study Group, n=30 | Case-Matched Controls, n=30 |
| --- | --- | --- |
| Intestinal Failure (IF) | 23 | 24 |
| -Short Bowel Syndrome (SBS) | 9 | 6 |
| -Crohn's Disease-Related Intestinal Failure (CDIF) | 4 | 4 |
| -Complication of Bariatric Surgery | 3 | 2 |
| -Gastroparesis | 3 | 4 |
| -Chronic Bowel Obstruction | 2 | 6 |
| -Celiac Disease | 1 | 1 |
| -Colitis | 1 | 1 |
| Enteric Fistula | 7 | 4 |
| Gastrointestinal Cancers | 3 | 5 |
| Pancreatic Cancer | 3 | 0 |
| Fallopian Tube Cancer | 1 | 1 |

Note. Principal diagnosis listed in the electronic medical record (EMR) for study patients and case-matched controls. Most HPN patients in the study and case-matched control groups had intestinal failure as their reason for therapy (23). Intrabdominal/pelvic cancer accounted for the remainder.

**Table 9.** Catheter characteristics of study patients and controls.

| Catheter Characteristics | Study Group, n=30 | Case-Matched Controls, n=30 |
| --- | --- | --- |
| Type: |  |  |
| - Infusion Port | 11 | 9 |
| - Tunneled Catheter | 10 | 5 |
| - PICC | 6 | 16 |
| - Groshong | 3 | 0 |
| Nursing Care |  |  |
| - Amerita | 25 | 7 |
| - Home Health Agency Nurses | 5 | 18 |
| - Clinic Staff | 0 | 5 |
| Dressings |  |  |
| - Bio-occlusive | 21 | 29 |
| - Hypoallergenic | 5 | 1 |

Note. Catheter type and care characteristics of study patients and case-matched controls. Most patients received catheter care on a weekly basis. Note: PICC = peripherally inserted central catheter.

**Table 10.** PN Orders Changes in study patients and controls.

| PN Order Changes | Study Group, n=30 | Case-Matched Controls, n=30 |
| --- | --- | --- |
| Macronutrients | 104 | 73 |
| Electrolytes | 173 | 178 |
| Micronutrients | 42 | 12 |
| Volume | 33 | 24 |
| Infusion Rate | 11 | 13 |
| Total PN Order Changes | 373 | 309 |

Note. PN orders for macronutrients were modified more often in study patients than in case-matched controls. Electrolytes, PN volume and duration were modified similarly among the study patients and case-matched controls. Micronutrients were modified in study patients more often than case-matched controls. The total number of PN formula changes were 20% higher in study patients than in case-matched controls. Note: PN = parenteral nutrition.

**Table 11.**
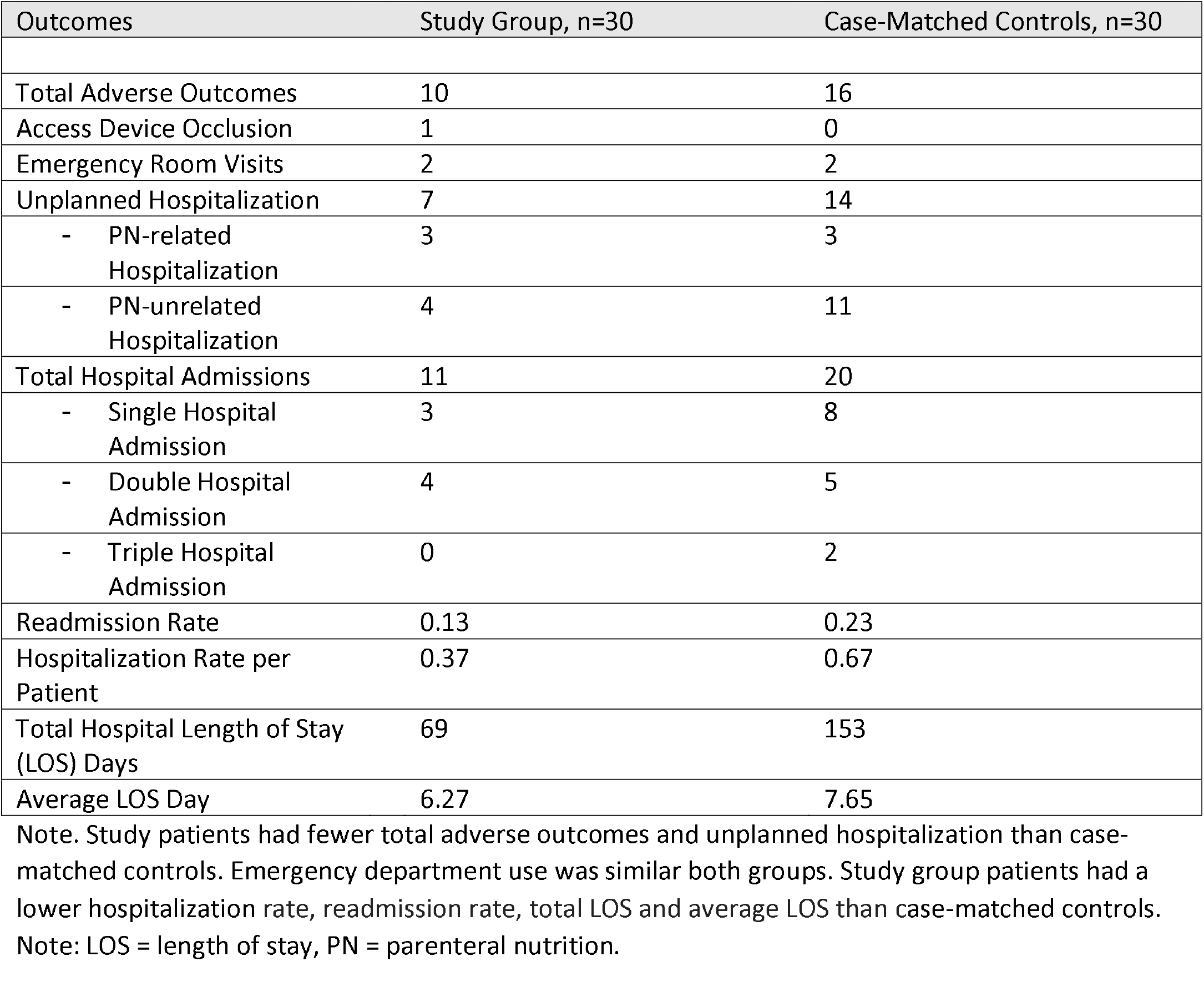
Adverse Outcomes in study patients and controls.

**Table 12.**
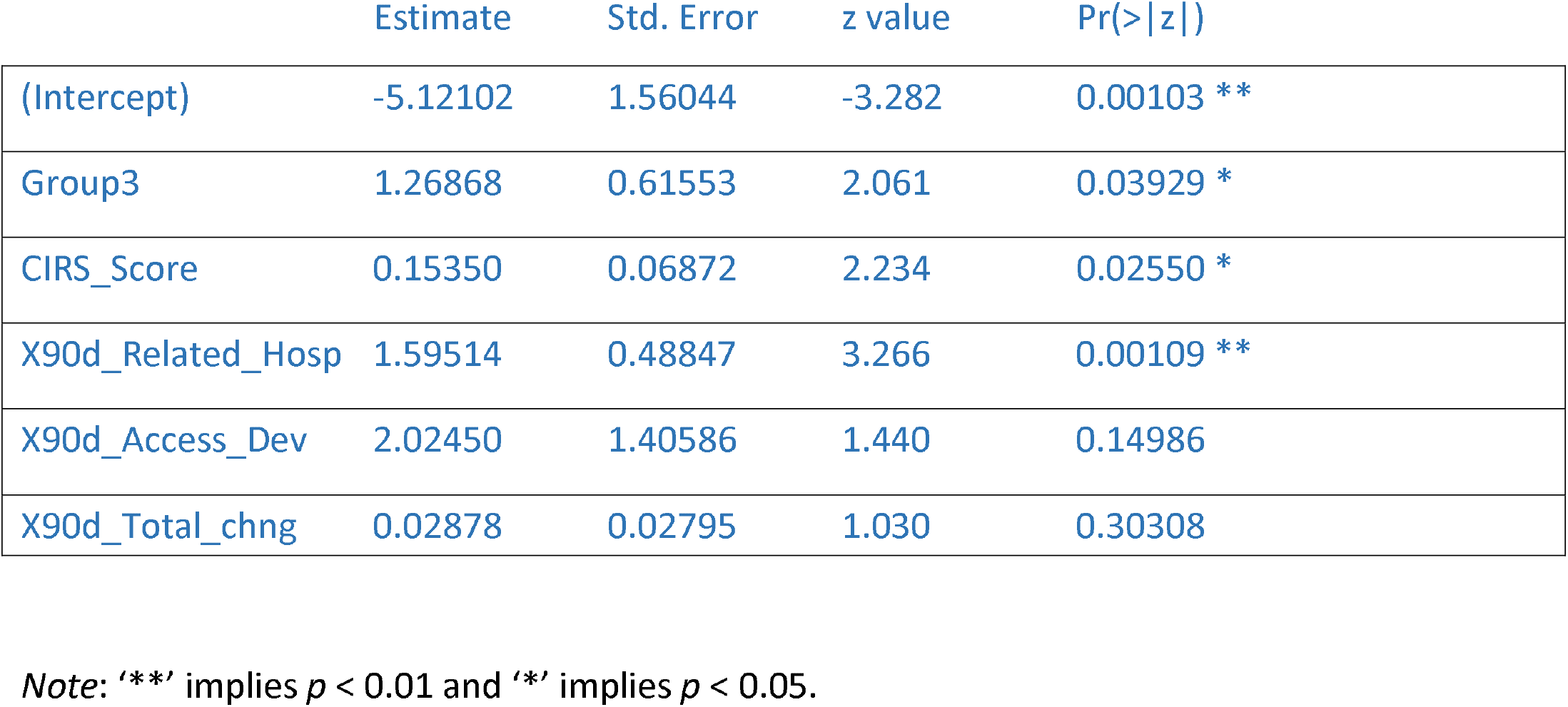
Results of modeling outcome variable, rate of adverse events per 90-day period, based on zero-inflated count data.

Figure 2 shows the Spearman correlation among these variables. Additionally, we chose group (with control group as reference group), CIRS Score as other predictors. The results of negative binomial regression are displayed in Table 5. Based on the results, group, CIRS Score, and 90-day hospitalizations, were significant predictors of 90-day (adverse) events at 5% alpha level. Specifically, rate of 90-day adverse events was significantly higher (3.56 times) for the control group than that for study group, given all other predictors are in the model. Likewise, for a unit change in CIRS Score, the percent change in expected incident rate of total 90-day events is by 16.6% (i.e., e^(0.15350) - 1), given all other predictors are in the model. For a unit change in 90-day unplanned hospitalization - related to therapy (X90d_Related_Hosp), the percent change in expected incident rate of total 90-day events is by 393% (i.e., e^(1.59541) – 1), given all other predictors are in the model.

**Figure 2.**
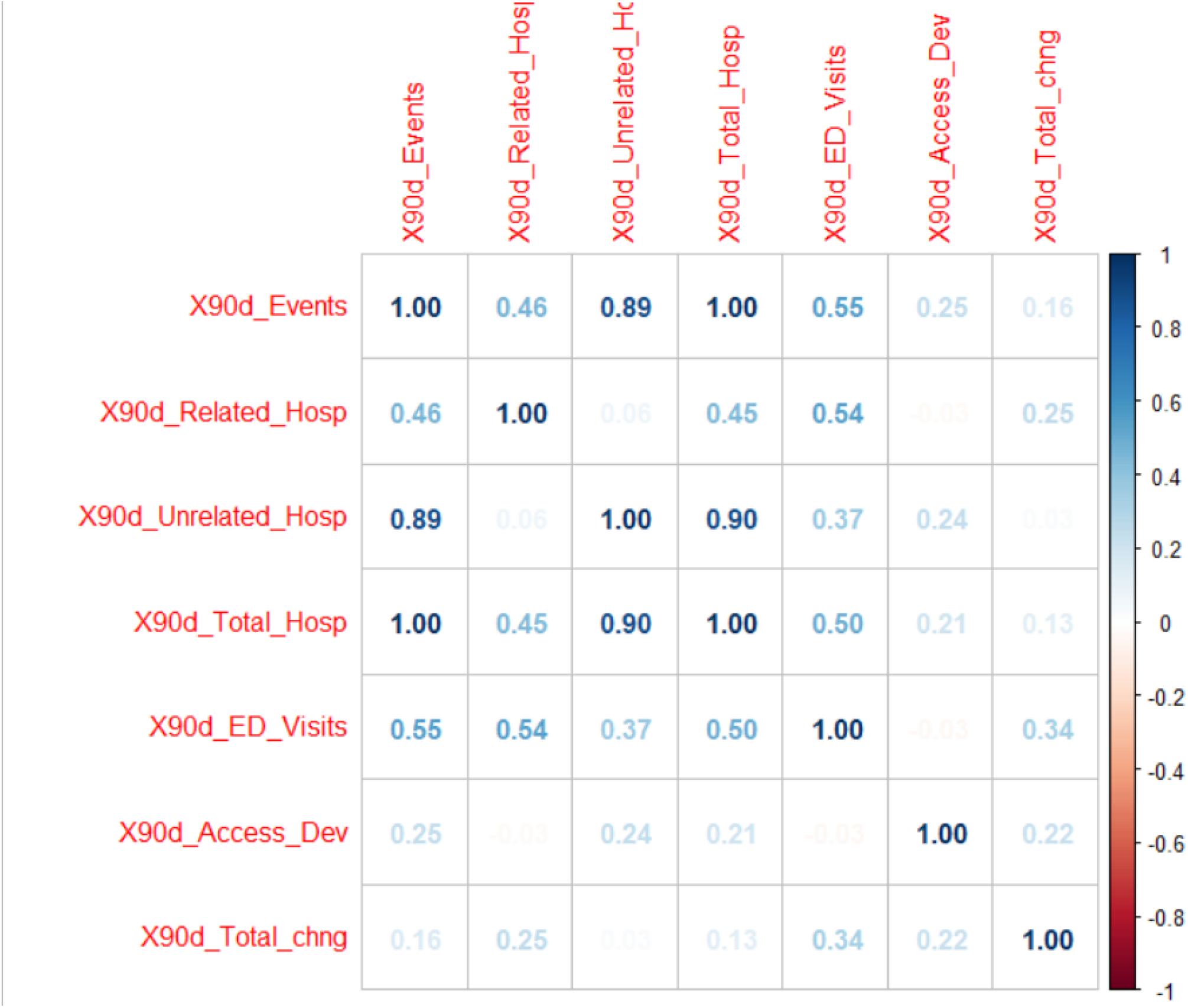
Correlation plot of Spearman correlation to show relationships of seven count variables in the data file (with only Study and Case Control Group patients). X90d_Events = total 90-day adverse events, X90d_Related_Hosp = 90-day hospitalizations related to therapy, X90d_Unrelated_Hosp = 90-day hospitalizations unrelated to therapy, X90d_Total_Hosp = 90-day total number of hospitalizations, X90d_ED_Visits = 90-day emergency department visits, X90d_Access_Dev = 90-day access device complications.

**Figure 3.**
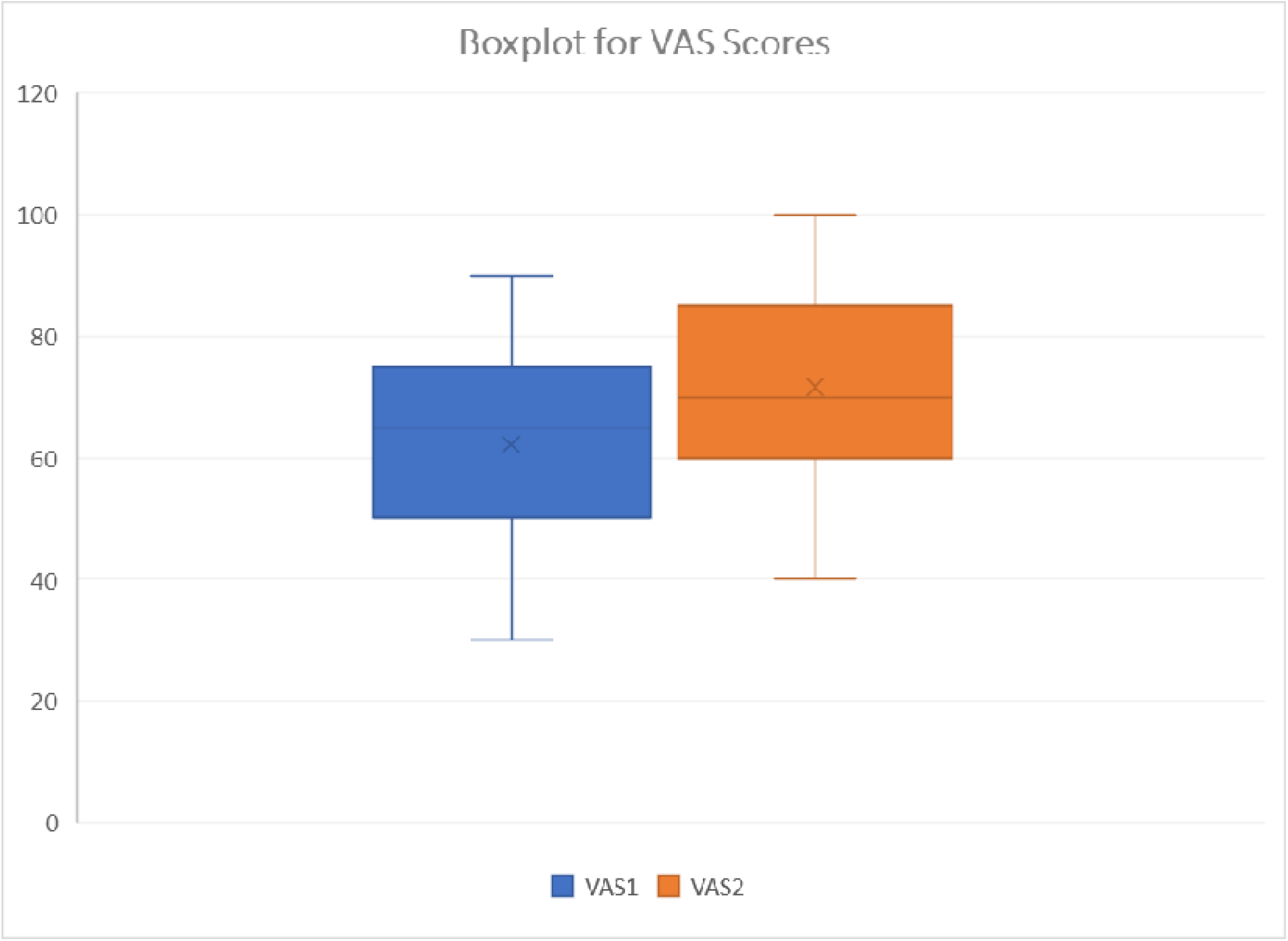
Boxplot representation of QOL EQ-5D 3L VAS scores before and after phase 3. QOL = quality of life, EQ-5D 3L = Euroquol 5 dimension, 3 level quality of life index, VAS = visual analog scale

## Discussion

This study demonstrated that a PNE-led, multidisciplinary team produced quality improvements for long term HPN patients. Patients who received MNST interventions had improved nutritional status, fewer adverse outcomes, hospitalizations, hospital LOS and better quality-of-life index scores.

MNST recommendations produced HPN quality improvement through compliance to standards of care and attention to PN formula adaptation based on physical findings and biochemical parameters. PN macronutrient changes resulted in clinical improvement, measured by weight gain and increased BMI. Improved biochemical parameters were seen in electrolyte balance, liver function, micronutrient balance and triglyceride levels. Study patients had significant improvement in QOL as measured by the EQ-5D-3L VAS.

An important aspect of the QIP-PN program was the difference of recommendations made and accepted by the treating physicians. The majority of HPN treating physicians were not nutrition specialists. Therefore, they relied upon the MNST to suggest changes that complied with standards or were indicated by HPN monitoring. The MNST made 157 recommendations for HPN management and 373 suggestions for formula changes during intervention. The vast majority (87.2%) of these recommendations were accepted by the treating physicians. Fewer recommendations for compliance were required if the patient’s physician was a PNE. Physicians who were not PNEs were more willing to accept the MNST’s recommendations.

We utilized the EQ-5D-3L QOL index to monitor study patients before and after intervention. Although other QOL indexes have been utilized in HPN, we found that the EQ-5D-3L index was better suited for the study because of its simplicity and patient acceptance. Other HPN QOL indexes employ up to 20 measured parameters compared to 5 for the EQ-5D-3L. In addition, we found that the VAS score provided a useful single point of reference for the patients overall perceived condition.

The study made use of a CIRS measure of multi-morbidity. We previously reviewed this and hypothesized its application in HPN research (35). To our knowledge this is the first real world application of the approach in the HPN population. CIRS scores documented the complexity of HPN patients with a moderately high value in both the study patients and controls.

### Total Hospitalizations and Length of Stay (LOS)

We demonstrated fewer hospitalizations in study patients versus controls. The average length of stay was 1.38 days shorter in study patients and there were 84 fewer hospital days among study patients than controls. The study also demonstrated a reduced rate of readmission to the hospital for study patients. None of the study patients was readmitted more than twice, while two of the control patients were admitted three times. The reductions in hospitalization, LOS and re-hospitalization would be expected to have a significant impact in the overall cost of care for HPN patients.

### Adverse Outcomes

When the phase 3 study patients were compared to the 90-day control patients for monitored adverse outcomes, statistically significant differences were found between the groups in total outcomes and unplanned hospitalizations. CIRS Score, and 90-day therapy-related hospitalizations were significant predictors of total outcomes at the 5% alpha level.

### Limitations

Our study group was small (30 patients and 30 controls) and the duration of the intervention period was short (60-90 days). Considering these limitations, additional research should be performed with a larger number of patients and longer timeframes of monitoring to affirm our favorable results.

## Conclusion

This study demonstrated that a PNE-led, multidisciplinary nutrition support team produced measurable improvements in the care of long term HPN patients. The MNST made numerous recommendations for HPN management, most of which were accepted by the treating physicians. The QIP-PN process improved patients’ self-assessed overall health, while reducing adverse outcomes, re-hospitalization and hospital LOS. If extended to the entire population of long-term HPN patients, we believe that MNST input would be expected to have a significant impact on the quality and cost of HPN care.

## Data Availability

All data produced in the present study are available upon reasonable request to the authors

## Brief Summary

We demonstrated that a physician nutrition expert (PNE)-led, multidisciplinary nutrition support team) MNST improved the care of long term HPN patients. MNST input can improve the quality and cost of HPN care.

## Statement of authors’ contributions to manuscript

Dr. Rothkopf designed the study and served as principal investigator.

Dr. Pant reviewed the data and performed statistical analyses.

Ms. Brown collected data and participated on MNST weekly patient rounds and discussions.

Ms. Haselhorst collected data and participated on MNST weekly patient rounds and discussions.

Ms. Gagliardotto participated on MNST weekly patient rounds and discussions and communicated recommendations to patients’ treating physicians.

Ms. Tallman participated on MNST weekly patient rounds and discussions and communicated recommendations to patients’ treating physicians.

Ms. Stevenson participated on MNST weekly patient rounds and discussions and communicated recommendations to patients’ treating physicians.

Mr. DePalma participated on MNST weekly patient rounds and discussions.

Mr. Saracco participated on MNST weekly patient rounds and discussions. Mr. Rosenberg collected data for phase 1b of the study.

Dr. Proudan served on the Study Oversight and Safety Committee.

Dr. Shareef served on the Study Oversight and Safety Committee.

Dr. Ayub served on the Study Oversight and Safety Committee.

